# Normative trajectories of extra-axial cerebrospinal fluid during childhood and adolescence defined in a clinically-acquired MRI dataset

**DOI:** 10.1101/2024.09.24.24314251

**Authors:** Ayan S. Mandal, Lena Dorfschmidt, Jenna M. Schabdach, Margaret Gardner, Benjamin E. Yerys, Richard A. I. Bethlehem, Susan Sotardi, M. Katherine Henry, Joanne N. Wood, Barbara H. Chaiyachati, Aaron Alexander-Bloch, Jakob Seidlitz

## Abstract

**Background:** Extra-axial cerebrospinal fluid (eaCSF) refers to the CSF in the subarachnoid spaces that surrounds the brain parenchyma. Benign enlargement of the subarachnoid space (BESS), a condition marked by increased eaCSF thickness, has been associated with macrocephaly and may be associated with subdural collections. However, diagnosis of BESS is complicated by the lack of age-specific normative data which hinders rigorous investigation of its clinical associations. Growth charts of eaCSF could shed light on normal CSF dynamics while also providing a normative benchmark to assist the diagnosis of BESS and other associated conditions.

**Methods:** We accessed clinically-acquired T1w MRI scans from 1226 pediatric patients to form a clinical control cohort. Nine scans from subjects with a diagnosis of BESS from a board-certified pediatric neuroradiologist were also reviewed. SynthSeg was used to segment each T1w scan into various tissue types, including eaCSF. Growth charts of eaCSF were modeled using the clinical control cohort. The confirmed BESS cases were then benchmarked against these charts to test the performance of eaCSF growth charts.

**Results:** eaCSF thickness varied nonlinearly with age, steadily decreasing from birth to two years, then trending upwards in early adolescence. Seven of the nine patients with a clinical diagnosis of BESS were above the 97.5^th^ percentile for their age for at least one eaCSF measure. Centile scores were able to distinguish BESS cases from controls with an area under curve (AUC) greater than 0.95.

**Discussion:** eaCSF thickness evolves in a dynamic pattern throughout childhood and adolescence. Patients with BESS can be differentiated from clinical controls using computational measurements of eaCSF thickness paired with normative modeling. Our findings demonstrate the feasibility of computational extraction of eaCSF with a potential point of clinical relevance, delineation of BESS diagnosis. Enhanced understanding of normative eaCSF is critical in further investigations its clinical associations.

## Introduction

Cerebrospinal fluid (CSF) plays a key role in the developing brain, namely in supporting neural stem cells, facilitating the distribution of cardinal signaling molecules, and clearing potential toxins.^1^ CSF is produced from blood in the choroid plexus, circulates through the ventricles, populates the basal cisterns and subarachnoid space, and is finally absorbed back into the venous circulation via the arachnoid granulations.^2^ Excess CSF volume, or hydrocephalus, can result from a blockage in the ventricular system (i.e. obstructive or non-communicating hydrocephalus) or impaired absorption from the arachnoid granulations (i.e. communicating hydrocephalus). Hydrocephalus before cranial suture closure leads to bulging fontanels and macrocephaly, and afterward, increased intracranial pressure, either of which can severely impair appropriate motor and cognitive development in children.^3^

More subtle CSF elevations, specifically within the subarachnoid space surrounding the brain parenchyma (i.e. extra-axial CSF; henceforth eaCSF), have traditionally been thought to be benign.^4,5^ However, more recent research has linked elevated eaCSF to multiple pathologies. Three studies in independent cohorts have linked increased eaCSF volume to the development of autism.^6–8^ Shen and colleagues demonstrated that findings of increased eaCSF volume can be detected as early as six months in infants later diagnosed with autism and that the increased eaCSF volume persists up to three years of age.^7^ eaCSF volume was further demonstrated to be associated with lower verbal ability and greater sleep disturbances.^9^

Benign enlargement of the subarachnoid space (BESS) is a common cause of macrocephaly and is marked by elevated eaCSF that typically self-resolves within 1-2 years of age.^4,5,10^ Current definition for BESS is largely based upon two-dimensional manual measurements of craniocortical, sinocortical, and interhemispheric width, with normal values derived from ultrasound based studies.^11,12^ While typically benign, the condition has been proposed to be associated with subdural collections with reported prevalence of such collections in BESS varying widely in the literature, from 0 to 42%, as recently reviewed.^13–17^ Clarity in this possible association would benefit from rigorous research built upon a standardized definition of BESS based on current, normative curves, that reflect the potential dynamic nature of early brain development.

Here, we describe a novel, automated computational pipeline to measure eaCSF thickness and apply this to a large cohort of clinical controls with limited imaging pathology.^18^ We aimed to define the normative trajectory of eaCSF in early childhood and adolescence, to both shed light on CSF dynamics during early postnatal development as well as provide a normative benchmark to assist in future research on BESS and associated conditions.

## Methods

### Participants

We accessed 2569 T1w scans from 1571 patients at the Children’s Hospital of Philadelphia (CHOP) as a part of the Scans with Limited Imaging Pathology (SLIP) cohort. The curation of this dataset is described in a prior publication from our group,^18^ though this study reports an updated cohort with more subjects. Briefly, a set of MRI scan sessions determined to lack clinically significant pathology, via review of signed reports from pediatric neuroradiologists, were retrieved from the CHOP Vender Neutral Archive. A team of clinical researchers also screened the radiology reports to remove scans suggestive of pathology using a grading process described in more detail in the Supplementary Materials. Notably, this process did not explicitly exclude scans where enlargement of the subarachnoid space or macrocephaly were mentioned but any mention of hemorrhage did result in exclusion.

For this study, we focused specifically on scans obtained using an approximately 1mm-istropic magnetization-prepared rapid acquisition gradient echo (MPRAGE) T1w sequence used for routine brain MRIs. We limited our study to subjects between the ages of 0 and 20 years, which reduced the number of scans and patients to 2560 and 1566 respectively. After subsequent exclusions of subjects described below and in Figure 1, we had a final sample of 1205 subjects described in Table 1.

**Figure 1.**
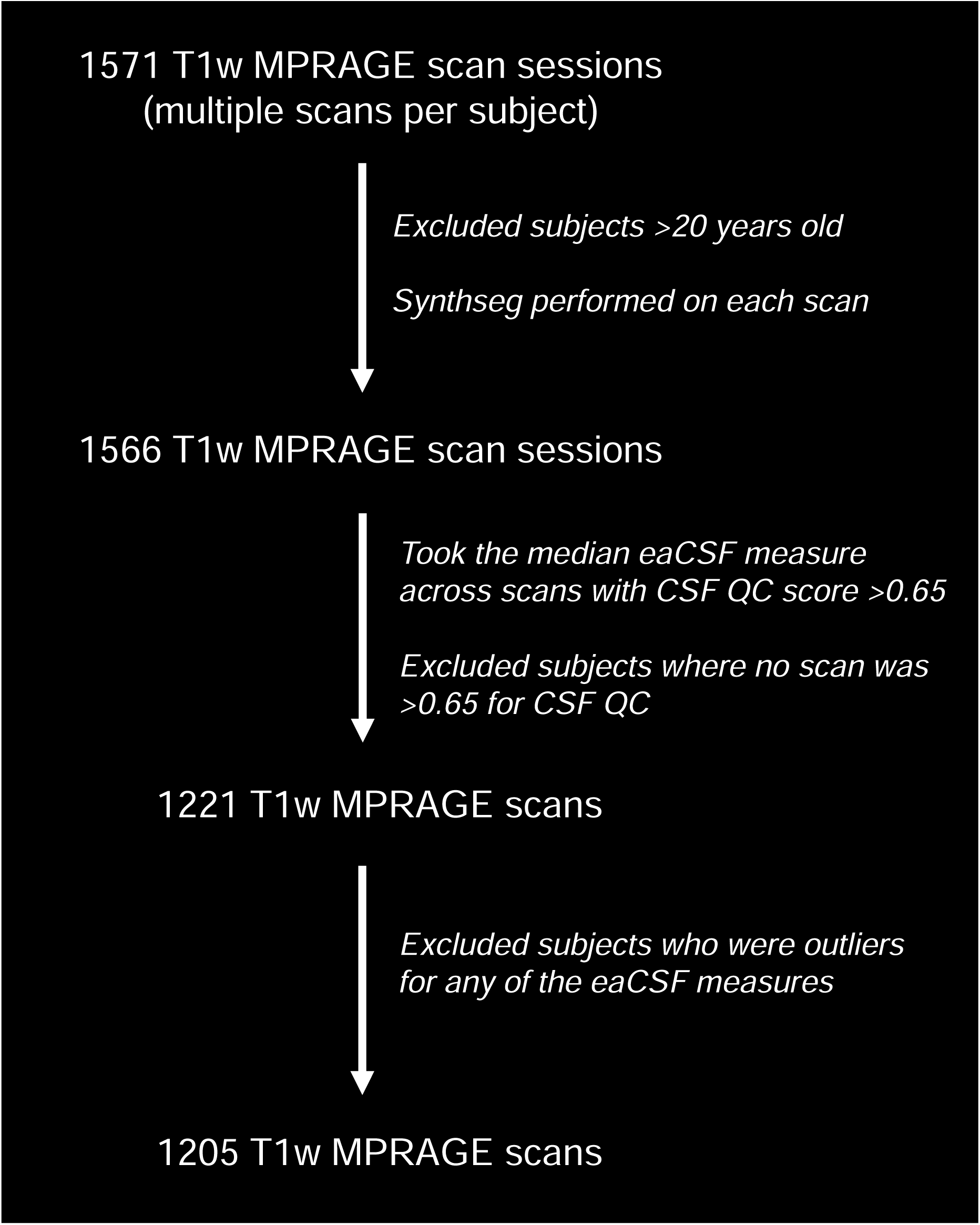
Flowchart outlining the exclusion criteria leading to the SLIP (i.e. clinical control) cohort.

**Table 1.** Demographics of patients in the final SLIP and BESS-CAP cohorts. Age is corrected for differences in gestational age at birth, if known. Standard deviations are included in parentheses. Full demographic information was not available for the BESS-CAP cohort due to privacy restrictions.

|  | **SLIP Cohort** |  |  | **BESS-CAP Cohort** |  |  |
| --- | --- | --- | --- | --- | --- | --- |
|  | **Total** | **Male** | **Female** | **Total** | **Male** | **Female** |
| **No. of Individuals Scanned** | 1205 | 586 | 619 | 9 | 8 | 1 |
| ***Age group (years)*** |  |  |  |  |  |  |
| **0 - 2** | 135 | 73 | 62 | 9 | 8 | 1 |
| **2 – 5** | 183 | 110 | 73 | 0 | 0 | 0 |
| **5 - 10** | 200 | 105 | 95 | 0 | 0 | 0 |
| **10 – 13** | 141 | 77 | 64 | 0 | 0 | 0 |
| **13 - 18** | 494 | 200 | 294 | 0 | 0 | 0 |
| **18 - 20** | 52 | 21 | 31 | 0 | 0 | 0 |
| ***Gestational age at birth (weeks)*** |  |  |  |  |  |  |
| **<34** | 20 | 8 | 12 | NA | NA | NA |
| **34 - 36** | 45 | 27 | 18 | NA | NA | NA |
| **37 - 40** | 353 | 176 | 177 | NA | NA | NA |
| **>40** | 40 | 20 | 20 | NA | NA | NA |
| **NA** | 747 | 355 | 392 | 9 | 8 | 1 |
| ***Race*** |  |  |  |  |  |  |
| **American Indian or Alaskan Native** | 1 | 0 | 1 | NA | NA | NA |
| **Asian** | 44 | 22 | 22 | NA | NA | NA |
| **Black or African American** | 292 | 158 | 134 | NA | NA | NA |
| **Multiple races** | 32 | 15 | 17 | NA | NA | NA |
| **Other** | 135 | 64 | 71 | NA | NA | NA |
| **White** | 694 | 325 | 369 | NA | NA | NA |
| **Unknown** | 1 | 0 | 1 | NA | NA | NA |
| **Refused** | 6 | 2 | 4 | NA | NA | NA |

In parallel, we accessed T1w scans from nine patients with clinical imaging read as diagnosis of BESS. This cohort of patients had an evaluation due to clinical concern for child physical abuse by Child Abuse Pediatrician (CAP) team and will henceforth be referred to as BESS-CAP. These anonymized scans underwent the same imaging pipeline, described below, as those in the SLIP cohort but were excluded from normative modeling. Scans from example subjects of similar age in the SLIP and BESS-CAP cohorts respectively are displayed in Figure 2A and B. This study was reviewed by the CHOP institutional review board and was determined to be exempt from institutional review board oversight as secondary analyses of preexisting clinical data.

**Figure 2.**
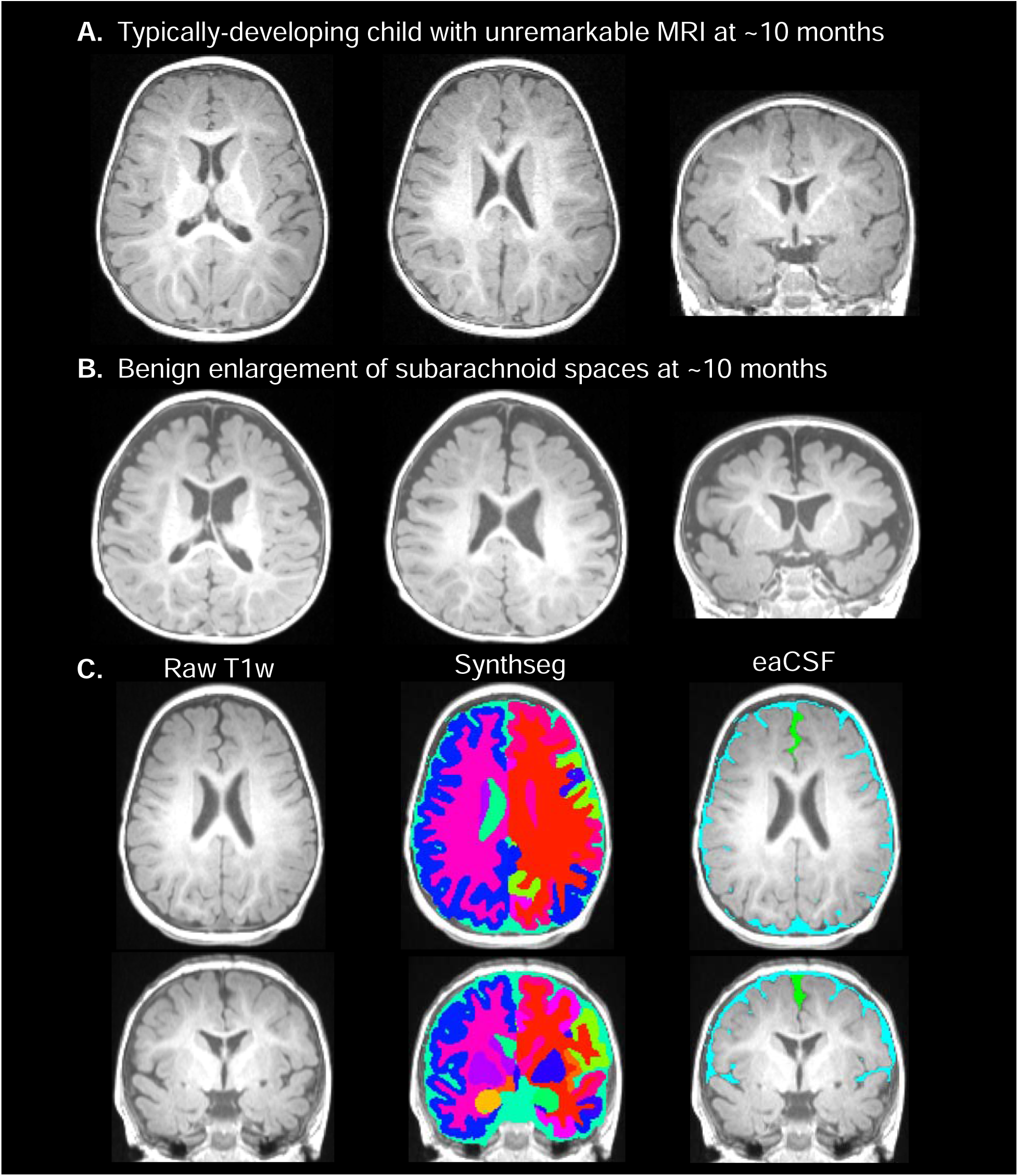
Quantitative approach to the assessment of eaCSF. (A) Typically developing patient in the clinical control cohort. (B) Patient diagnosed with benign enlargement of the subarachnoid space. (C) Pipeline for extracting eaCSF measures. SynthSeg segments T1w scans by tissue compartment, from which eaCSF in the subarachnoid space (light blue + green) and interhemispheric fissure (green) can be estimated.

### Tissue segmentation

All T1w scans were analyzed using SynthSeg, a segmentation algorithm based on a convolutional neural network approach that outputs volumetric masks for a comprehensive set of intracranial tissue compartments, including eaCSF.^19^ SynthSeg was run with the robust flag and the SynthSeg 2.0 version was used (SynthSeg+). Quality control metrics outputted by SynthSeg+ were used to screen out scans for which the algorithm could not perform optimally due to motion degradation or other issues (discussed in more detail below).

### Computing measures of eaCSF thickness

The component of eaCSF most pertinent to BESS is known to surround the frontal and temporal gyri, while the CSF in the basal cisterns is less relevant.^4^ Consistent with prior studies, we focused on supratentorial eaCSF by removing the portion of the eaCSF segmentation mask inferior to a plane linearly registered to run parallel with the anterior commissure.^7,8^ For the calculation of eaCSF within the interhemispheric fissure, we further isolated the mask to cover the anterior component of the interhemispheric fissure using manually-drawn templates registered to each T1w scan’s native space (Figure 2C).

Thickness was computed for each eaCSF mask using built-in functions in AFNI.^20^ We applied the @measure_bb_thick function to the total, supratentorial eaCSF mask (blue + green in Figure 2C) to compute the thickness of the subarachnoid space. This algorithm computes a thickness at each voxel by building the largest possible sphere centered at that voxel that still fits within the mask, then outputting the size of that largest sphere. To compute an automated measure that would correspond more closely to a manually derived measure in the axial plane, we applied the @measure_erosion function to the interhemispheric eaCSF mask (green in Figure 2C) to compute the thickness of the interhemispheric fissure. This algorithm computes a thickness at each voxel by iteratively performing a simple morphological erosion to the mask, removing voxels at the edge of the mask. The number of iterations required before a given voxel is eliminated is equal to the thickness at that voxel. The median thickness across all voxels was reported as the thickness of the corresponding eaCSF measure.

In addition to directly modeling subarachnoid space thickness and interhemispheric fissure thickness, we also computed models in which these measures were normalized using total intracranial volume. Total intracranial volume was also estimated using SynthSeg+ and produces a measure that correlates well with clinical measurements of head circumference (Supplementary Figure 1).

For each given imaging measure, scans with poor quality control (QC) scores for the tissue type of interest were screened out. Specifically, for all measures of eaCSF, scans with a quality control score for CSF lower than 0.65 were excluded following a previously validated procedure.^19^ An average of the QC scores from all tissue compartments was used to threshold scans for total intracranial volume. Of the scans that passed QC for a given imaging measure for a given subject, the median of that measure across scans was taken to reflect the value for that subject.

### Outlier detection

Because the normative trajectories in this study were computed within a cohort of clinical controls, it was important to exclude scans with highly abnormal tissue volumes that may indicate pathology. On the other hand, it was also important to permit an appropriate degree of variance for each measure that is expected in clinical populations. We defined outliers as subjects for whom the absolute value of a modified Z-score exceeded 3.5 for a given imaging measure.

A modified Z score was calculated first by fitting a simple spline model with four degrees of freedom predicting a given imaging measure with age and sex as covariates. The residuals of this model were used as the numerator of the Z score, whereas the denominator was calculated by multiplying the median absolute deviation with a constant scale factor *k* of 1.4826 (assuming normally distributed data).

Subjects who were outliers for at least one of the eaCSF measures were excluded from all normative modeling. Subjects who were outliers for total intracranial volume were excluded from the specific analyses involving that measure but not the other models. For subjects who were under two years of age and outliers for at least one of the eaCSF measures, we accessed the radiology report of the corresponding scan to determine if there was any mention of enlarged subarachnoid spaces consistent with BESS. While these subjects were not included in the normative modeling, we used the procedure outlined below to calculate their centile scores, or percentiles corresponding to the percentage of subjects of the same age and sex below a given subject.

### Growth chart modeling

Growth charts for each eaCSF measure were fit with generalized additive models for location, scale, and shape (GAMLSS), a flexible, distribution-based regression approach that models mean, variance, and higher statistical moments.^19^ This process involved selecting an appropriate distributional family that captured the data most parsimoniously over the studied age range, which resulted in the selection of a customized version of the generalized gamma distribution family with robust mean and variance (GGalt; https://github.com/brainchart/Lifespan), as it most consistently produced the lowest Akaike Information Criterion (AIC) across phenotypes (Supplementary Figure 2). Age and sex were included as predictor variables (fixed effects) in each model whereas scanner ID was included as a random effect. Only data from the SLIP (i.e. clinical control) cohort were used to build the growth chart models.

For each phenotype of interest, the normative trajectories, variance, and rate of growth were plotted including both the median across the age range as well as the 2.5^th^ percentile and 97.5^th^ percentile, each segregated by sex. Since a fixed effect of sex would not account for interaction effects of age and sex, we also performed the growth chart modeling on males and females separately (see Supplementary Figure 3 and 4). Further, to assess potential effects of gestational age at birth, we performed an additional sensitivity analysis, where we used a subset of the data for whom gestational age at birth was known (see Supplementary Figure 5).

Once the GAMLSS models were built, we computed percentiles for each eaCSF measure for both the subjects in the clinical control cohort and the BESS-CAP cohort. When scanner ID was not available, a percentile was calculated using a model in which the random effect was removed (which affected one BESS subject). Percentiles for each eaCSF measure were used to plot receiver operating characteristics (ROC) curves to test their ability to distinguish between BESS-CAP cases and clinical controls. Finally, we accessed the radiology reports for subjects in the clinical control cohort with at least one eaCSF measure greater than the 97.5th percentile to determine how commonly the extra-axial spaces were mentioned in the original read amongst these subjects.

## Results

### Normative trajectories of eaCSF in the clinical control cohort

In the clinical control cohort, measures of eaCSF thickness evolved nonlinearly with age, declining over the first years of life before stabilizing in early childhood (Figure 3). While measures of interhemispheric fissure thickness plateau for the rest of the studied age range, subarachnoid space thickness begins to increase around six years of age, corresponding with the age at which cortical gray matter volume starts to decline (Figure 3B and D).^18^ The degree of increase in subarachnoid space thickness after six years of age is diminished when accounting for the effects of total intracranial volume (TIV), yet still positive (Figure 3B and D). Median trajectories for subarachnoid space and interhemispheric fissure thickness showed moderate correspondence with analogous measures assessed using ultrasound.^21^

**Figure 3.**
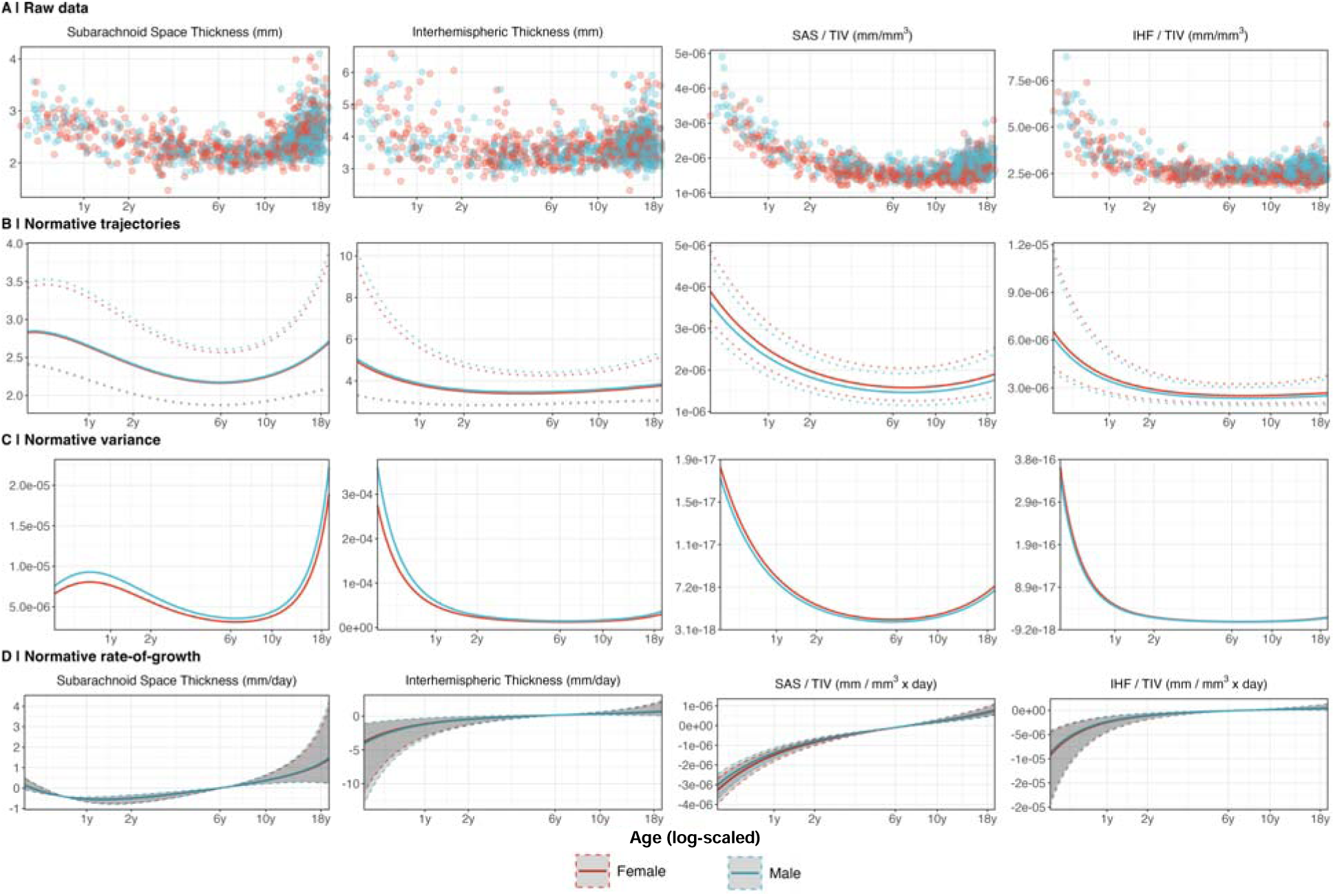
Growth charting of eaCSF measures. (A) Raw data for each eaCSF measure plotted. (B) Median (i.e. 50^th^ percentile) trajectories for each eaCSF measure plotted, with the dotted lines corresponding to the 2.5^th^ and 97.5^th^ percentile for males and females respectively. (C) Variance for males and females for each eaCSF measure across the age range. (D) Normative rate-of-growth for each eaCSF measure, with the dotted lines corresponding to the 2.5^th^ and 97.5^th^ percentile for males and females respectively. Abbreviations: SAS = subarachnoid space; IHF = interhemispheric fissure; TIV = total intracranial volume.

These patterns generally reproduce when producing separate models for patients of different sex (Supplementary Figures 3 and 4) and when limiting the analysis to subjects for whom gestational age at birth is available (Supplementary Figure 5). Interestingly, while median trajectories of subarachnoid space and interhemispheric fissure thickness do not vary substantially by sex, variance in eaCSF amongst males, especially during the age range when BESS is diagnosed, is greater than in females (Figure 3C).

### Growth charts of eaCSF separate BESS cases from clinical controls

Using the growth chart models built from clinical control data, we calculated centile scores for eaCSF thickness among BESS-CAP cases and clinical controls. BESS-CAP cases generally appeared around the 97.5^th^ percentile lines for each growth chart (Figure 4). We found that BESS-CAP cases could be reliably distinguished from clinical controls using these centiles scores (Figure 4), with each measure producing an ROC curve with an area under curve (AUC) greater than 0.95. Of the nine clinical BESS subjects, seven had at least one eaCSF measure >97.5^th^ percentile, with several being above this threshold for multiple measures (Table 2).

**Figure 4.**
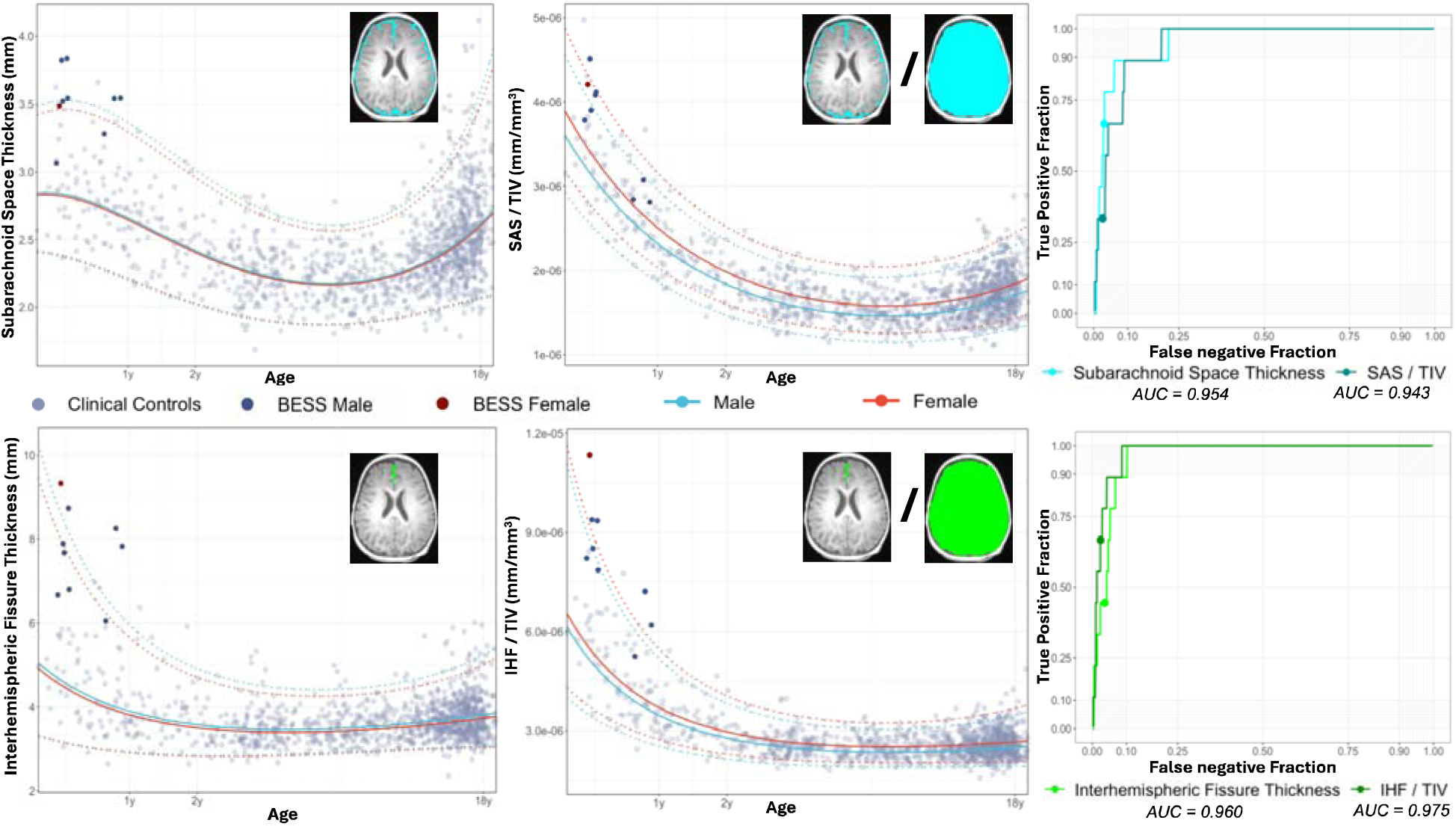
Growth charts of fully automated eaCSF measures separate BESS-CAP cases from controls. Plots in the top left, bottom left, top middle, bottom left, and bottom middle show growth charts for median subarachnoid space thickness (SAS), interhemispheric thickness (IHT), ratio of SAS and total intracranial volume (TIV), and ratio of IHF and TIV respectively, with BESS cases colored separately. Receiver operator curve (ROC) plots shown on the right demonstrating performance characteristics of using the derived percentiles from the eaCSF models in distinguishing between BESS-CAP cases and clinical controls. Solid circles in each ROC plot represent the true positive rate and false negative rate of each measure if using the 97.5^th^ percentile as the threshold for BESS-CAP versus control.

**Table 2.** Percentiles of eaCSF measures for the BESS-CAP cohort. Centile scores above the 97.5^th^ percentile are bolded. Age and gender are omitted due to privacy restrictions.

| *Subject* | *SAS* | *IHF* | *SAS / TIV* | *IHF / TIV* |
| --- | --- | --- | --- | --- |
| Subject 1 | **97.9%** | **99.1%** | 95.9% | **99.5%** |
| Subject 2 | **97.7%** | 92.9% | **99.2%** | 96.9% |
| Subject 3 | 93.9% | 95.2% | 80.3% | 91.8% |
| Subject 4 | 76.8% | 89.0% | 90.4% | 95.2% |
| Subject 5 | 97.4% | 95.7% | 96.5% | **97.6%** |
| Subject 6 | **99.5%** | **98.3%** | **99.0%** | **99.2%** |
| Subject 7 | **98.8%** | **99.7%** | 90.7% | **99.4%** |
| Subject 8 | **98.6%** | **99.7%** | 96.6% | **99.8%** |
| Subject 9 | **99.5%** | 96.2% | **99.8%** | **98.8%** |

Finally, we examined the radiology reports for subjects under age two in the SLIP cohort who had abnormally elevated eaCSF to determine how commonly abnormalities of the extra-axial spaces were mentioned in the original read for these subjects. These subjects were identified in two ways. Six subjects were identified as outliers for at least one eaCSF measure based on a modified Z-score greater than 3.5 calculated using a simple spline model and were accordingly excluded from the GAMLSS normative modeling. All six of these subjects were outliers for interhemispheric fissure thickness alone. Ten subjects were not outliers and were thus included in the normative modeling, but they were identified to have at least one eaCSF measure greater than the 97.5^th^ percentile.

Extra-axial spaces were mentioned to be abnormal in seven out of the 16 patients who met either criteria, with BESS having been mentioned explicitly for three of these subjects (Table 3). Curiously, the six subjects who were outliers for interhemispheric fissure thickness based on the simple spline model had lower centile scores than one might expect given a modified Z-score of 3.5 or greater. The discrepancy likely results from the assumption of constant variance in the simple spline model which is inaccurate especially during the first two years of life for this measure (Figure 3C).

**Table 3.** Percentiles of eaCSF thickness for subjects in the SLIP cohort under the age of two with at least one abnormally elevated eaCSF measure. Centile scores above the 97.5^th^ percentile are bolded.

| *Sex* | *SAS* | *IHF* | *SAS /*<br>*TIV* | *IHF*<br>*/*<br>*TIV* | *Clinical*<br>*indication* | *Radiology report*<br>*findings* |
| --- | --- | --- | --- | --- | --- | --- |
| *Subjects identified as outliers for at least one eaCSF measure* |  |  |  |  |  |  |
| Male | 92.2% | **99.8**<br>**%** | 91.5<br>% | **99.9**<br>**%** | NF1 | Unremarkable |
| Male | 92.0% | 96.4% | 81.9<br>% | 96.2<br>% | Motor<br>developmental<br>delay | **Concern for BESS**<br>**mentioned** |
| Male | **98.8%** | **99.7**<br>**%** | **98.4**<br>**%** | **99.7**<br>**%** | New onset<br>seizures | **Concern for BESS**<br>**mentioned** |
| Male | 88.7% | 89.7% | 75.1<br>% | 90.3<br>% | Suspicious<br>fracture | Thin subdural fluid<br>collection over frontal<br>lobes |
| Femal<br>e | 91.5% | **98.2**<br>**%** | 53.4<br>% | 92.6<br>% | Lesion on head<br>CT; new onset<br>seizures | Lesion resolved; **stable**<br>**bifrontal subarachnoid**<br>**spaces** |
| Male | 82.7% | 97.1% | 38.4<br>% | 93.5<br>% | Seizure | **Prominence of CSF**<br>**spaces over frontal**<br>**lobes** |
| *Subjects identified as >97.5<sup>th</sup> percentile for at least one eaCSF measure* |  |  |  |  |  |  |
| Femal<br>e | **99.4%** | **98.5**<br>**%** | 73.5<br>% | 93.1<br>% | Developmental<br>delay | Unremarkable |
| Male | **97.6%** | **99.7**<br>**%** | 96.1<br>% | **99.7**<br>**%** | Sturge Weber<br>syndrome.<br>Hemangioma on<br>right<br>forehead/scalp/fa<br>ce | **Minimal prominence**<br>**of the extra-axial**<br>**spaces over the**<br>**bilateral frontal lobes** |
| Femal<br>e | **97.8%** | 82.8% | **99.7**<br>**%** | 93.9<br>% | Eyelid swelling | Unremarkable |
| Male | 84.0% | 76.5% | **98.1**<br>**%** | 92.8<br>% | Developmental<br>delay | Unremarkable |
| Femal<br>e | **98.1%** | **99.5**<br>**%** | 85.8<br>% | 98.5<br>% | Seizure; CT<br>showed<br>questionable<br>hemorrhage | **Concern for BESS**<br>**mentioned.** No<br>evidence of hemorrhage. |
| Male | 95.3% | 97.4% | **99.0%** | **99.5%** | Extremity tremor and head titubation | Unremarkable |
| Female | 84.9% | 97.4% | 93.2% | **98.6%** | Skull lesion | Ovoid lesion is seen in the left posterior calvarium (but no intracranial mass lesions). Otherwise unremarkable. |
| Male | 84.4% | **98.1%** | 65.7% | 96.9% | Simple and complex febrile convulsions | Subtle T2 and FLAIR signal abnormalities of unclear significance. Otherwise unremarkable. |
| Female | 69.5% | **97.8%** | 67.6% | 95.2% | Monitor for lesion progression or new lesions; malignant neoplasm of eye | Unchanged T2 hypointense nodule in near globe. No focal brain lesion visible.<br>**Bifrontal extra-axial fluid spaces remain prominent.** |
| Female | **99.1%** | 61.6% | **99.8%** | 77.7% | Abnormal head ultrasound, history of one lifetime seizure | Unremarkable |

## Discussion

Here, we report the first growth charts of eaCSF, demonstrating that eaCSF thickness changes in a dynamic but relatively predictable way over the early lifespan. These findings offer insights into the timing of certain neurodevelopmental pathologies and show promise for the development of machine-assisted diagnostic tools for the evaluation of BESS.

### Normative trajectories of eaCSF

eaCSF is known to change dynamically as the brain parenchyma grows and shrinks over the lifespan,^22^ though the exact timing of these changes has never been fully characterized. We demonstrate that eaCSF declines sharply over the first two years of life before stabilizing in early childhood. The cause of this decline is thought to reflect maturation of the arachnoid granulations which absorb CSF back into the venous circulation,^2,3^ though other mechanisms likely also play a role, such as closure of the cranial sutures forcing the CSF to reflux into the perivascular spaces and the brain parenchyma.^9^ Garic and colleagues recently demonstrated that elevated eaCSF during the first two years of life correlates with dilatation of the perivascular spaces at 24 months of age, which in turn was linked to an elevated rate of autism.^9^ Speculatively, it is possible that the association between elevated eaCSF and the diagnosis of autism could relate to delayed maturation of the arachnoid granulations affecting the clearance of CSF from brain parenchyma.

One future direction would be to further investigate the associations between sex and eaCSF dynamics. We find that while the median trajectory of eaCSF thickness is similar between males and females, variance is greater in males, especially over the course of the early developmental period. Pronounced differences in variance in males compared to females is also observed when the interhemispheric fissure thickness is measured via ultrasound during this same developmental period.^21^ Sex differences in variance become slightly less pronounced later in the lifespan during childhood and adolescence, but both the median trajectory and the variance of subarachnoid space thickness rises in general during adolescence perhaps reflecting pubertal effects.

The diagnosis of BESS largely relies on static and manually derived measures of craniocaudal width, sinocortical width, and/or interhemispheric distance,^11,12^ yet our study demonstrates that these measures change dynamically over early infancy. We show that a computationally-derived measure of eaCSF thickness can be benchmarked against and a growth chart reference such that individuals are compared to the reference sample accounting for their age and sex. Centile scores calculated from eaCSF growth charts performed extremely well in distinguishing a small sample of confirmed BESS patients from clinical controls. Furthermore, subsequent analyses of the radiology reports of scans from the clinical control group revealed that many of the subjects with high eaCSF thickness were also suspected to have BESS. Taken together, these findings provide strong evidence eaCSF growth charts to assist in future research on BESS and, pending further validation studies, play a role in the diagnosis of BESS in clinical contexts. A compelling future direction would be to understand if computationally-derived measurements of the eaCSF in combination with growth charts could be useful for future research assessing the clinical associations of altered eaCSF dynamics.

### Limitations

A potential limitation of the study is that the population used to derive normative trajectories is composed of patients who were scanned at a hospital due to some clinical indication, and therefore cannot necessarily be considered healthy. In some ways, this is a virtue since this sample is more representative of a patient population than subjects in research studies whose differences from the general population have been well-documented.^23^ Nevertheless, the normative trajectories reported here should be interpreted as a growth reference as opposed to a growth standard,^24^ and this is reflected in the fact that some subjects in the clinical control cohort have evidence of BESS. Reassuringly, we note that prior work from our group has demonstrated that growth charts of other neuroimaging phenotypes derived from this same clinical cohort mirror those derived from controls in prospective research studies.^18^

Another limitation is the limited number of CAP-identified BESS subjects included in this study. The relatively small number of clinical reports denoting BESS mitigates our opportunity to use growth charts to better understand this condition. For example, it is unclear whether BESS patients have some degree of ventricular enlargement, a question that should be addressed by future research. Child abuse evaluations as source of clinical BESS cases may also introduce selection bias. Evaluation of eaCSF growth charts within rigorous study designs would be necessary to clarify relevance of these eaCSF measurements with any association with subdural collections, if present. In addition, while automated segmentation of eaCSF was gated for homogeneity that would exclude other components such as blood, it is possible that volumes extracted from BESS case subjects may include other extra-axial collections. Reassuringly, cases without subdural collections were identified with BESS within clinical reads among the SLIP cohort. Further segmentation review is warranted in more cases with multi-compartment collections. Although similar in range to manually-derived measures of eaCSF,^21^ the growth charts presented herein should not be used to benchmark manually-derived caliper measurements. Larger samples and further clinical validation are required before eaCSF charts should be used in clinical practice. On the other hand, our clinical control cohort represents the largest ever reported sample for which eaCSF has been measured, and therefore helps to characterize the development of this clinically relevant phenotype.

In summary, we present growth charts of eaCSF over the early lifespan that shed light on the timing and demographics of multiple clinical conditions related to eaCSF elevation. This work lays the foundation for further studies that could lead to clinical applications of brain growth charts in the evaluation of BESS, a common cause of macrocephaly.

## Data Availability

All data produced in the present study are available upon reasonable request to the authors, except for that which may compromise patient privacy

## Supplementary Information

### Supplementary Methods

#### SLIP Radiology Report Grading

The goal of SLIP cohort curation was to select scans that could be analyzed using standard image processing pipelines extracting brain structure volumes and thicknesses without issue. In doing so, it was necessary to exclude scans with significant imaging pathology that may interfere with the accuracy of these pipelines. A team of researchers were trained to read radiology reports from potential SLIP subjects and assign one of the three grades below:

<u>Rating 0</u>: Suspect serious imaging pathology or unusable scan

<u>Rating 1</u>: Unclear whether the reported findings should be interpreted as pathology

<u>Rating 2</u>: No reason to suspect imaging pathology

Graders were trained to assign a rating of 0 to scans that mentioned terms suggestive of severe pathology, including brain surgery, neoplasm, stroke, hemorrhage, aneurysm, herniation, atrophy, or severe motion or hardware-related (e.g. dental/orthodontic) artifacts. A grade of 1 was assigned to scans that described findings which could reflect significant brain pathology based on context, for example macrocephaly, microcephaly, optic neuritis, focal cortical dysplasia, or ventriculomegaly. A grade of 2 was given to scans that did not mention any of the above and were reported to be “unremarkable”. Grades were averaged across two raters, and scans where the averaged score was greater than or equal to 1 were included in the SLIP cohort. Any major disputes between graders were addressed in regularly scheduled meetings and discussed with one of the board-certified physicians on the team if certain issues recurred.

**Supplementary Figure 1.**
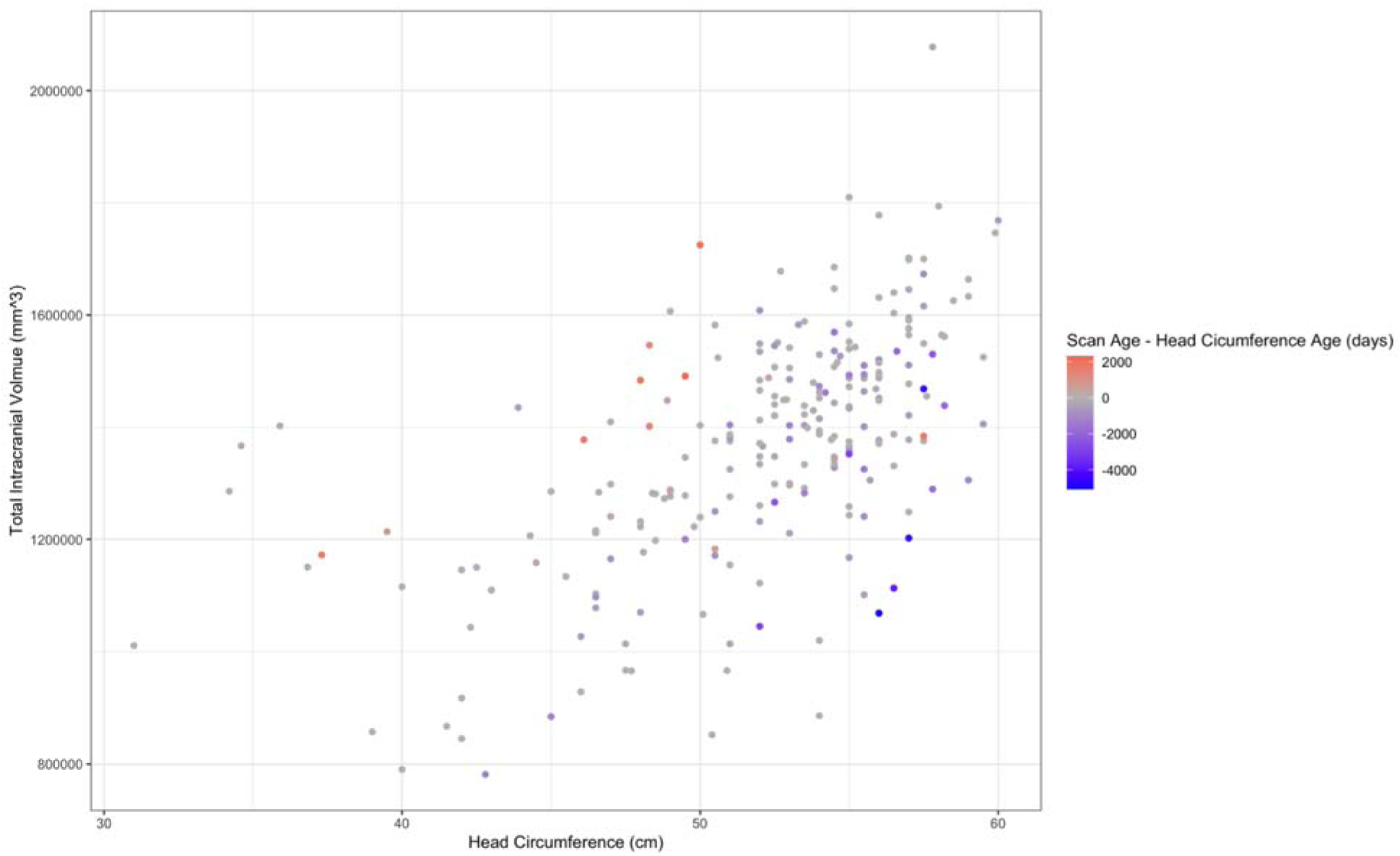
Relationship between total intracranial volume via MRI and head circumference measured at a similar timepoint. Total intracranial volume was correlated with manually measured head circumference for a subset of SLIP subjects for whom head circumference data were available (n = 236; ρ = 0.608; P < 2.2 × 10-16). Head circumference was generally measured on a different day from scan date, and datapoints are colored based on the discrepancies between these dates. The correlation increased when data were excluded to subjects whose scan and head circumference measurements were no more than 100 days apart (n = 128; ρ = 0.742; P < 2.2 × 10-16).

**Supplementary Figure 2.**
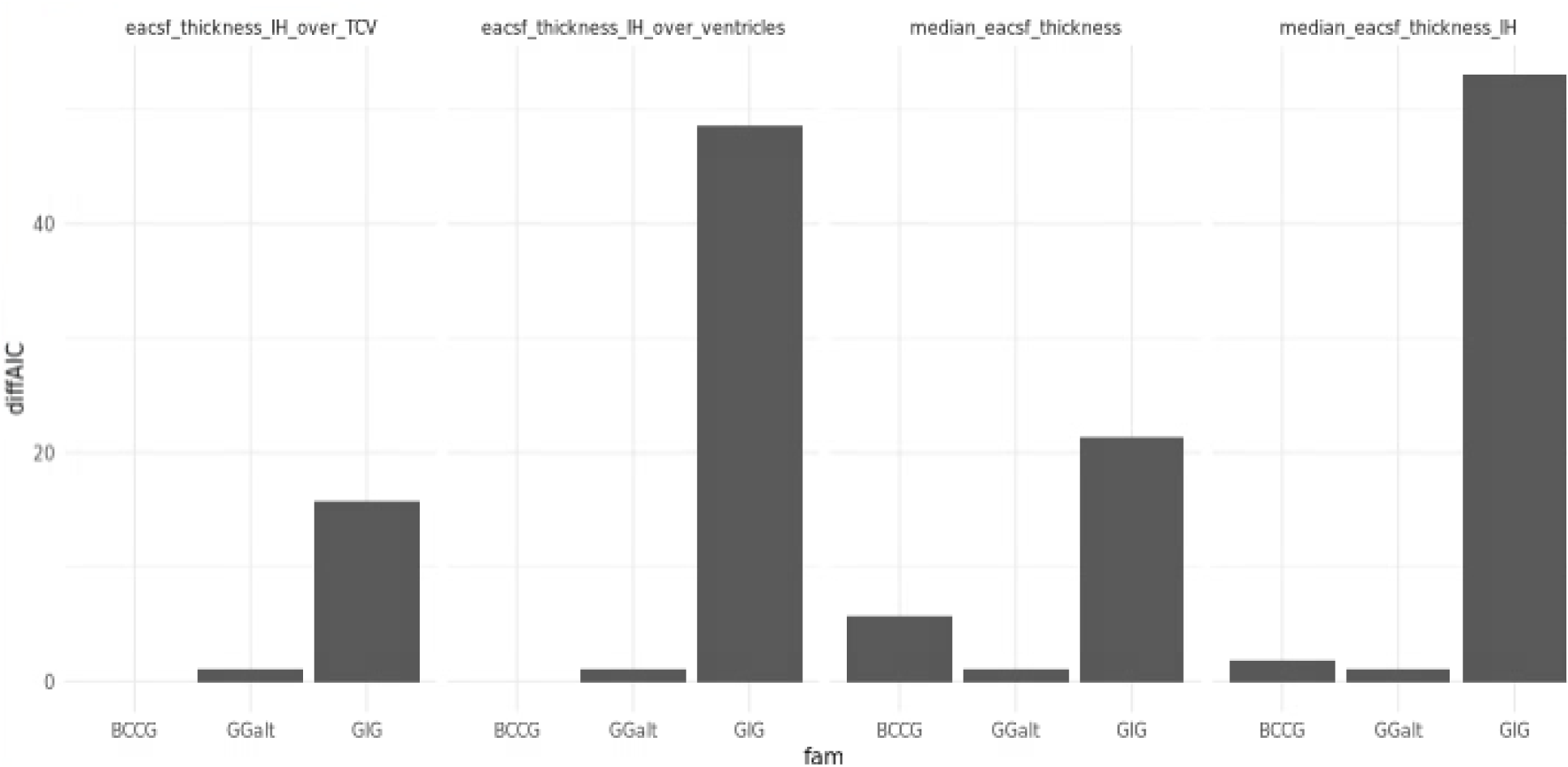
Family selection for GAMLSS models based on difference in AIC. Based on these results, the GGalt family was selected as the distributional family that best fit the data most consistently.

**Supplementary Figure 3.**
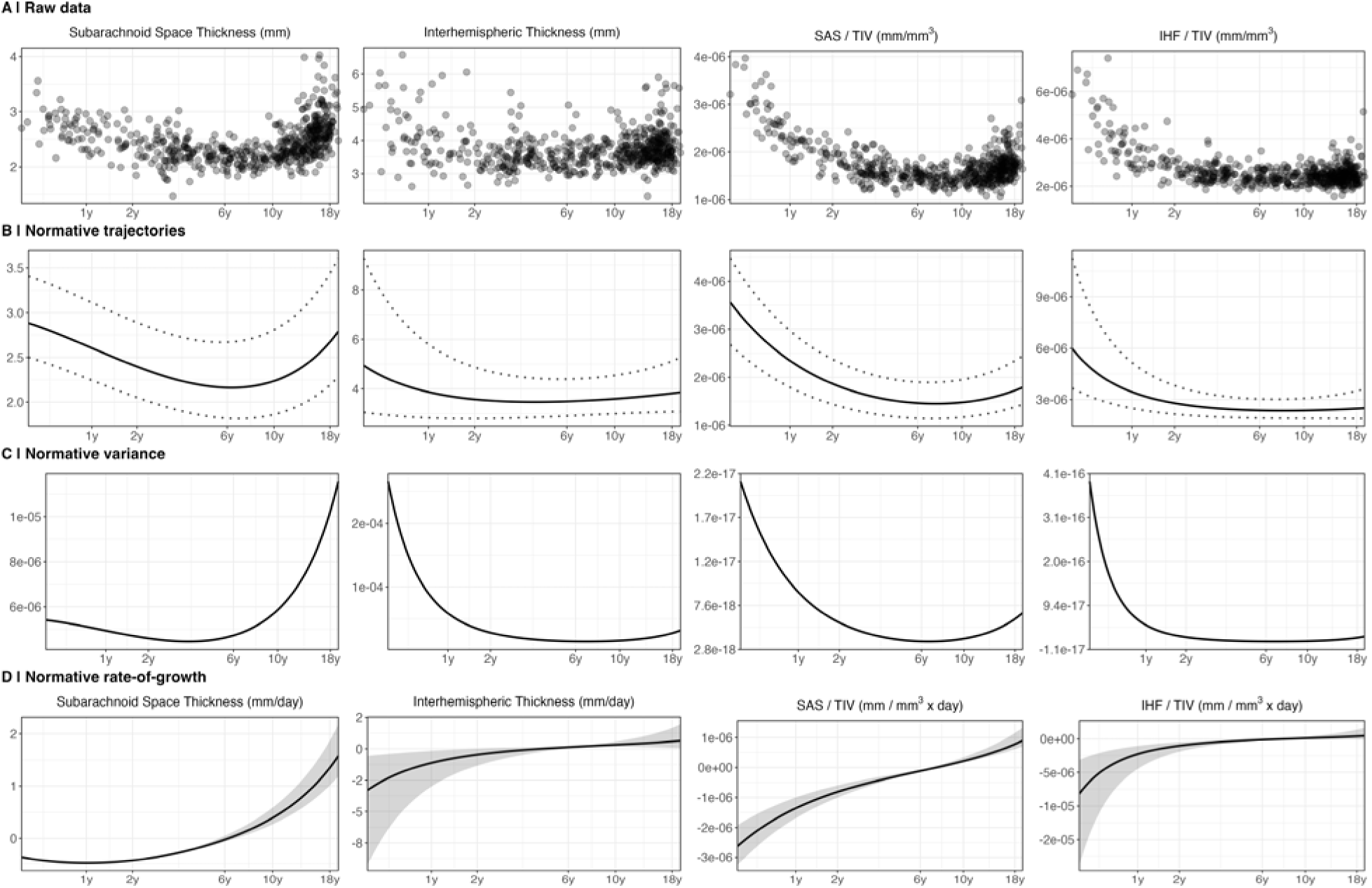
Sensitivity analysis of growth charts of eaCSF measures using only male patients.

**Supplementary Figure 4.**
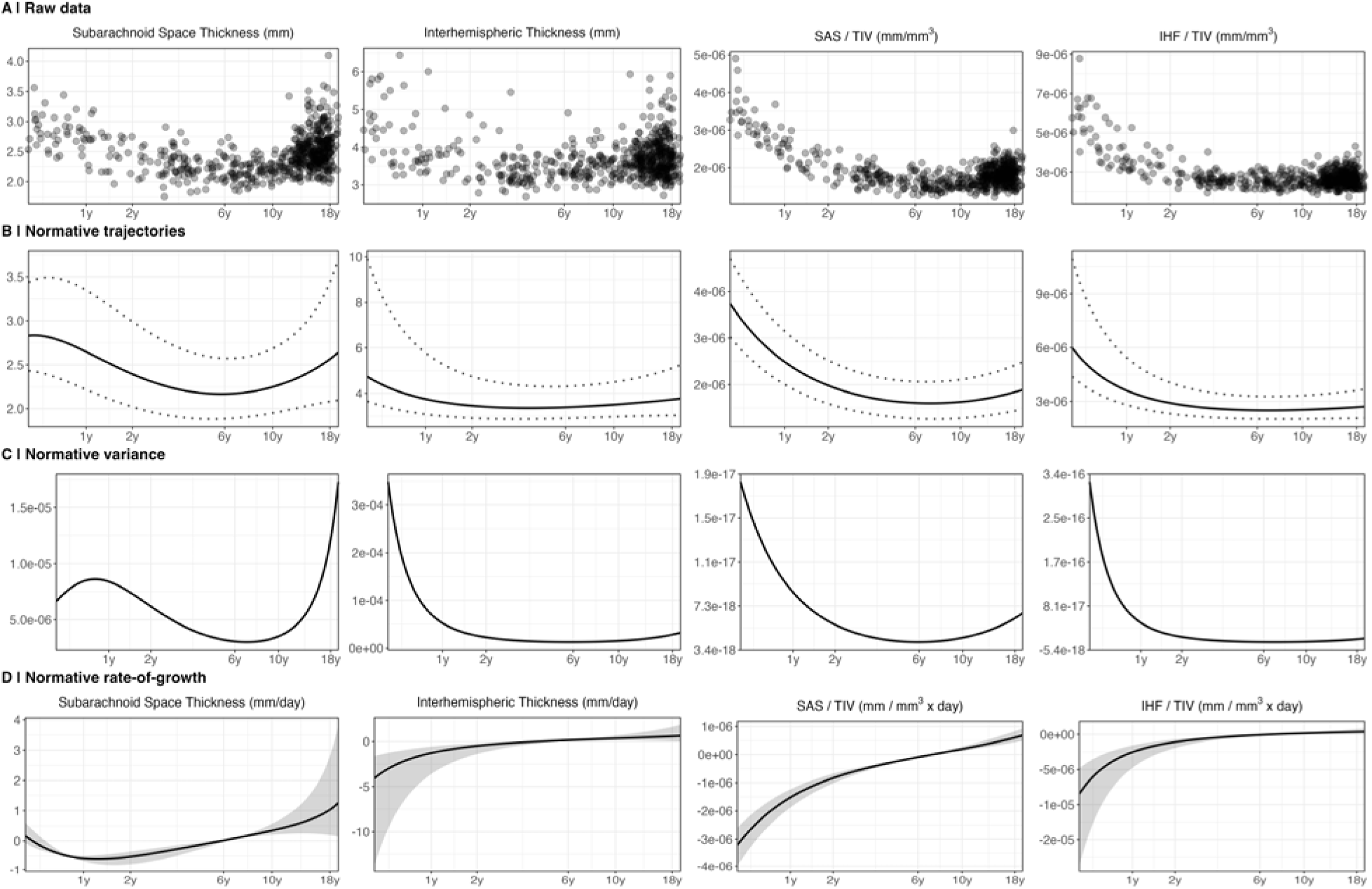
Sensitivity analysis of growth charts of eaCSF measures using only female patients.

**Supplementary Figure 5.**
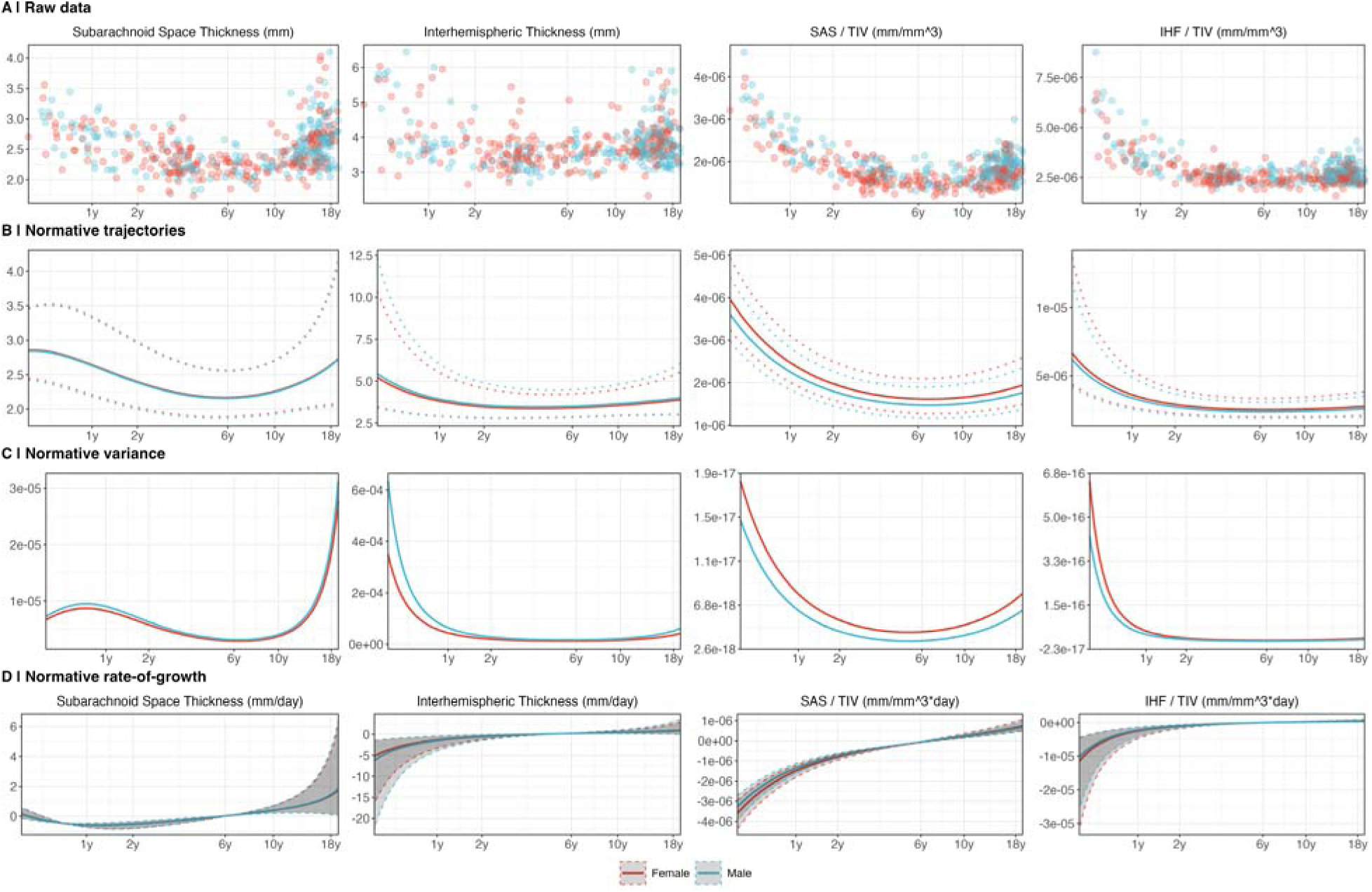
Sensitivity analysis of growth charts of eaCSF measures using only patients for whom gestational age at birth is available.

